# Endometrium receptivity and the dose-related effects of Acupuncture therapies in infertile women: a protocol for systematic review and meta analysis

**DOI:** 10.1101/2021.04.02.21254309

**Authors:** Xiaoyan Zheng, Liying Liu, Hang Zhou, Hongmei Yang, Fangge Wang, Jie Yang

## Abstract

**Introduction:** The aim of the systematic review (SR) is to evaluate the efficacy of Acupuncture in endometrium receptivity(ER) of infertile women and find out dose-related between Acupuncture and ER improving.

**Methods and analysis:** We will search four English databases: PubMed, Embase, Cochrane Library, Web of Science, and five Chinese databases: SinoMed (formerly Chinese Biomedical Database), CNKI (Chinese National Knowledge Infrastructure), Wanfang Data, and China Biomedical Literature Database (CBM), China Science Journal Database (VIP database) from inception to February 2021 in English and Chinese. Also, we will manually retrieve other resources, including reference lists of identified publications, conference articles, and grey literature. All clinical randomized controlled trials related to Acupuncture for endometrium receptivity of infertile women will be included. Two review authors will perform all research selection, data extraction, and research quality assessment. According to suggestions, data will be synthesized in a fixed-effect model, or random effect model due to the heterogeneity test. The primary outcomes include ER (endometrial thickness and endometrial hemodynamic parameters) and clinical pregnancy rate(CPR). Secondary outcomes include a dose of the interventions(the starting time of Acupuncture, the duration, and frequency of Acupuncture sessions), and adverse events will be assessed. We will use the statistical package (RevMan5.4.0) provided by The Cochrane Collaboration to analyze data. The quality of evidence will be assessed by using the Grading of Recommendations Assessment, Development, and Evaluation (GRADE) framework.

**Ethics and dissemination:** Since this article does not contain patient personal information, ethical approval is not required. The contract is distributed by a peer-review

**Systematic review registration:** PROSPERO, CRD42020206790

**Strengths and limitations of this study:** This study will be the first ever systematic review and meta-analysis in dose-related effects of Acupuncture therapies in treating endometrium receptivity.

The quality of evidence will be assessed by the Grading of Recommendations Assessment,Development, and Evaluation system.

Our research approach will only focus on the dosage of acupuncture interventions. Due to the diversity of included treatment plans, Traditional Chinese Medicine and Moxibustion will increase the heterogeneity of results.

We will only retrieve data from Chinese and English databases which could limit available data or result in language bias.

## 1. Introduction

In recent years, the incidence of infertility has been increasing year by year^[1,2]^. Infertility has gradually become the world’s third most common disease, second only to cardiovascular diseases and tumors.^[3]^.It has steadily become a problem waiting to be solved for couples of childbearing age.

Assisted reproductive technology(ART)represented by in vitro fertilization and embryo transfer (IVF-ET)has been rapidly developed, bringing hope to patients with infertility and realizing the dream of many families to breed offspring. IVF-ET pregnancy rate is becoming higher to 30-50% ^[4].^ Although IVF-ET has dramatically improved the fertilization rate and cleavage rate with advances in embryo culture technology, the current clinical pregnancy rate is still unsatisfy. Relevant studies have found that even if some patients have a normal ovarian function and no organic endometrial disease, under the premise of high-quality embryos in the IVF-ET cycle, the repeated implantation failure (RIF) still occurs. Embryo implantation is a complex biological process. Large-scale epidemiological studies^[5,6]^ have shown that a considerable number of women have repeated implantation failure after several embryo transfers. If embryo implantation fails for at least three consecutive treatment cycles during IVF treatment, and there are 1 or 2 high-quality embryos in each cycle^[7]^; or after 2-6 assisted reproduction treatment cycles, a total of ≥10 high-quality embryos, but still cannot be implanted which is called RlF ^[8,9]^. The reason has not, however, be exact. It is mostly considered from the abnormality of embryonic development potential, endometrial receptivity and synchronization of embryo and endometrium^[10]^, as well as an abnormal ovarian response^[11]^, inappropriate ovarian stimulation in IVF^[12]^, embryo transfer technology ^[13]^, etc. All of these factors will also lead to RlF. Among them, 2/3 of RIF is closely related to endometrial receptivity^[14]^. So improving endometrial receptivity is the key to enhance the IVF-ET pregnancy outcome.

Endometrial receptivity (ER) refers to the uterus’s ability to accept blastocyst implantation and make it grow. Psychoyos from France first proposed it in 1976. He also pointed out that the blastocyst development can only be successfully implanted if it changes synchronously with the endometrium. The endometrium can only accommodate embryo implantation for a short period, the “implantation window period”^[15].^The indicators of endometrial receptivity are ultrasound-endometrial thickness(the distance between the anterior and posterior walls of the endometrium, including the uterine cavity gap), endometrial Type (Gonen classification standard^[16]^: type A-trilinear or multilayered endometrium, strong echo in the outer and middle parts, hypoechoic or dark areas in the inner layer, and unmistakable linear echo in the uterine cavity; Type B-weak trilinear, isolated echo in the middle, inconspicuous echo in the middle of uterine cavity; Type C-mean strong echo, no intrauterine midline echo), blood flow (includes the uterine artery, endometrium, and endometrial blood flow^[17]^), morphology-pinocytosis, molecular-interleukin, integrin, leukemia inhibitory factor and so on. In the non-invasive detection method, the use of vaginal color Doppler ultrasound is a non-invasive, safe, and economical examination method. Therefore, endometrial thickness, endometrial shape, and uterine artery blood flow are common assessments of endometrial receptivity.

Modern medicine uses estrogen and progesterone to increase the thickness of the endometrium, supplement insufficient progesterone, and improve endocrine function; use low dose aspirin (LDA)^[18]^to improve the local blood supply of the uterus; Abnormal cavity; mechanical stimulation of the endometrium improves endometrial receptivity ^[19]^, oxytocin receptor antagonist atosiban reduces uterine tension and inhibits uterine contraction ^[20]^, low molecular heparin ^[21]^prevention Thrombus formation at the embryo implantation site and placenta attachment site during early pregnancy; intrauterine perfusion of granulocyte colony-stimulating factor G-CSF^[22]^ and other methods to improve IVF outcome. However, these methods are still not effective in improving embryo implantation. Stem cell transplantation or uterine transplantation is the most cutting-edge technology at present, but the relevant evidence for clinical application is still insufficient. Thus, there is no ideal medicine or methods can improve the problem of ER yet. Therefore, finding a simple, effective, and economical treatment has become an urgent need to increase the pregnancy rate.

Acupuncture has a history of thousands of years in the treatment of infertility in ancient China.And in 2002, Paulus of Germany and Professor Zhang Mingmin of Tongji University in China conducted a randomized controlled study of 160 cases of Acupuncture and a control group. Acupuncture was given 25 minutes before and after IVF-ET. Before embryo transfer, they used acupoints: Neiguan(PC6), Diji(SP8), Taichong(LR3), Baihui(DU20), and Guilai(ST29). After transplantation, they used acupoints: Zusanli(ST36), Sanyinjiao(SP6), Xuehai(SP10), and Hegu(LI4). It was found that Acupuncture significantly improved the clinical pregnancy rate ^[23]^. This research has caused significant repercussions in the field of reproductive medicine. Acupuncture, as a complementary alternative medicine(CAM) in ART, is mainly guided by the fundamental theories of traditional Chinese medicine (TCM)and selects relevant acupoints for treatment according to the differentiation of the meridians and collaterals. Relevant reports ^[24-28]^have proved that Acupuncture can improve endometrial thickness, uterine artery blood flow and increase clinical pregnancy rate to a certain extent. The research ^[29]^find out Acupuncture can regulate the endocrine system and the microenvironment of the ovary, increase the blood supply of the endometrium, and improve the receptivity of the endometrium.However, The scientific clinical significance of acupuncture is the subject of controversy. Emily Wing^[30]^find out there is no significant differences in the changes in reduction of endometrial and subendometrial vascularity between acupuncture group and placebo group.Placebo acupuncture and only twice treatment before and after embryo transfer are used in this trail.Therefore, the way of control group and the session of treatment or even the starting time of acupuncture maybe the key to evaluate the acupuncture efficiency.

We searched the published systematic reviews and meta-analyses^[31]^. As shown in **Table 1**, no meta-analysis included all kinds of patients with low ER,patients were undergoing IVF-ET only included. As we all know, patients with polycystic ovary syndrome will also have decrease endometrial receptivity due to endocrine disorders^[24]^. Besides, several new published trials were not be researched in the meta-analyses. ^[29]^Moreover, there is currently no feasible systematic review to evaluate treatment options, and the timing of Acupuncture intervention reported in the relevant literature is also different. The measurement of Acupuncture is also inconsistent.

**Table 1.** Summary of systematic reviews and meta-analyses of Acupuncture on the treatment of endometrium receptivity of infertility.

| Systematic review and meta-analysis | Number of the included study | Participants | Literature inclusion time | Outcome measures |
| --- | --- | --- | --- | --- |
| Zhong, et 2018<sup>[31]</sup> | n=13 | Infertility , low ER | 2012-2018 | ET, ER, RI, PI, S/D |
| Yang, et,2019<sup>[32]</sup> | n=12 | Infertility , ART, | 2012-2018 | ET, EP, RI, PI<br>Duration of Acupuncture |
| Hu et,2017<sup>[33]</sup> | n=8 | Low ER, ART | 2009-2015 | Percentage of pinocytosis<br>ET, EV, EP |

ER=endometrium receptivity, ET=endometrium thickness, EP=endometrium pattern, EV=endometrium volume, RI= resistive index, PI=pulse index, S/D=peak systolic velocity/ end-diastolic blood velocity, ART= assisted reproductive technology

Therefore, the purpose of this systematic review is to evaluate the effectiveness of Acupuncture in improving endometrial receptivity for all kinds of infertile woman with low ER and to screen its optimal Acupuncture plan (starting time and duration of Acupuncture intervention, optimal intervention measurement) to provide a more vigorous evidence-based clinical practice.

## 2. Methods and analysis

### 2.1. Design and registration of the review

The protocol has been registered on the prospective international register of systematic review (PROSPERO) (registration number is CRD42020206790). The protocol is designed strictly under the preferred reporting items of the systematic review and meta-analysis protocol (PRISMA-P).^[35]^The PRISMA Guidelines and the Cochrane Handbook will be used for the studies we evaluate for inclusion. Also, bias risk analysis and heterogeneity analysis will be used in our research. Subgroup analysis and sensitivity analysis will be further carried out.

### 2.2. Inclusion criteria

#### 2.2.1. Type of study

Randomized controlled clinical trials (RCT)will be searched. Any other type of literature will be excluded in Chinese and English due to the language limitation of our researchers.

#### 2.2.2. Types of participants

We include infertile women patients with low endometrial receptivity. Regardless of race, educational level, source of cases, and cause of illness. Also, repeated IVF failures will be included. Participants with other serious diseases, such as heart disease, liver disease, kidney disease, or cancer (especially ovarian and breast cancer) will be excluded from the trial.

#### 2.2.3. Diagnostic criteria

① Infertility: a patient who had unprotected, regular sexual life for one year but without conception, whose husband has average semen quality and shape, including primary infertility and secondary infertility. (World Health Organization (WHO) in 2002)

② Endometrium receptivity: evaluated by ultrasound includes endometrial thickness, endometrial pattern, endometrial hemodynamic parameters,such as resistance index (RI), pulse index (PI), endometrial vascular index (vascularization index, VI), flow index (FI), etc.

③ Thin endometrium: The endometrial thickness has not yet reached a consensus, but related studies^[36] [37]^ have found the endometrium thickness <7mm in the middle luteal phase (after ovulation) of 6∼10 days, the pregnancy rate is significantly reduced. So in this SR, the thin endometrium is when the follicle is mature, the endometrial thickness is < 7.0 mm. The uterus is regular in shape; ART has been or has not been performed. Informed consent has been signed.

#### 2.2.4. Types of interventions

The intervention measures should take Acupuncture(or transcutaneous acupoint electrical stimulation, TESA) or Acupuncture combined with other methods to treat female infertility with low ER. In contrast, the control group was treated with non-Acupuncture therapy, blank control group, or placebo Acupuncture (such as sham Acupuncture).

#### 2.2.5. Types of outcomes

In this systematic review, we will analyze embryo transfer rate, clinical pregnancy rate(CRP), endometrial thickness, endometrial pattern, helical arterial blood flow index including resistive index (RI), pulse index (PI), peak systolic velocity/end-diastolic blood velocity (S/D) to evaluate the efficacy of Acupuncture. Also the duration and starting time will be extracted to assess the relationship between dosage of Acupuncture and efficacy.

### 2.3. Data sources and search methods

#### 2.3.1. Electronic searches

We will search four English databases: PubMed, Embase, Cochrane Library, Web of Science, and five Chinese databases: SinoMed (formerly Chinese Biomedical Database), CNKI (Chinese National Knowledge Infrastructure), Wanfang Data, and China Biomedical Literature Database (CBM), China Science Journal Database (VIP database) from inception to July 2020. The search strategy will be based on the guidance of the Cochrane handbook. The language was limited to English and Chinese. The strategy of search for PubMed is shown in **Table 2**. We will use this search strategy in several other databases.

**Table 2.**
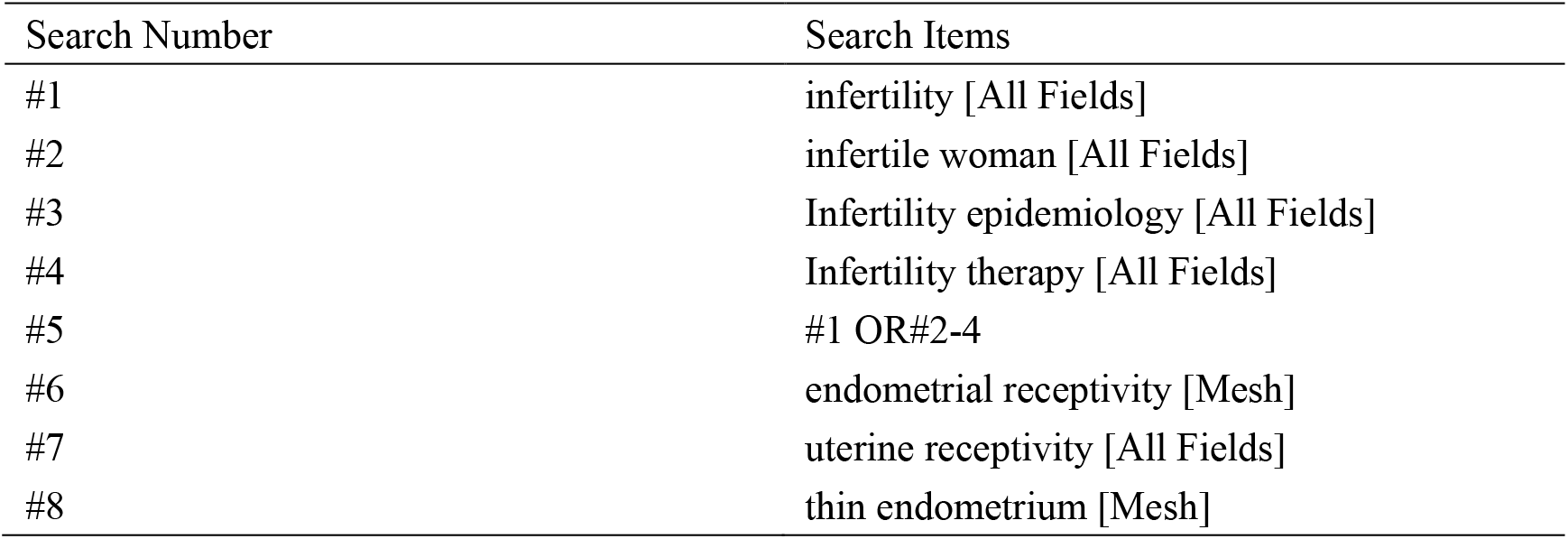
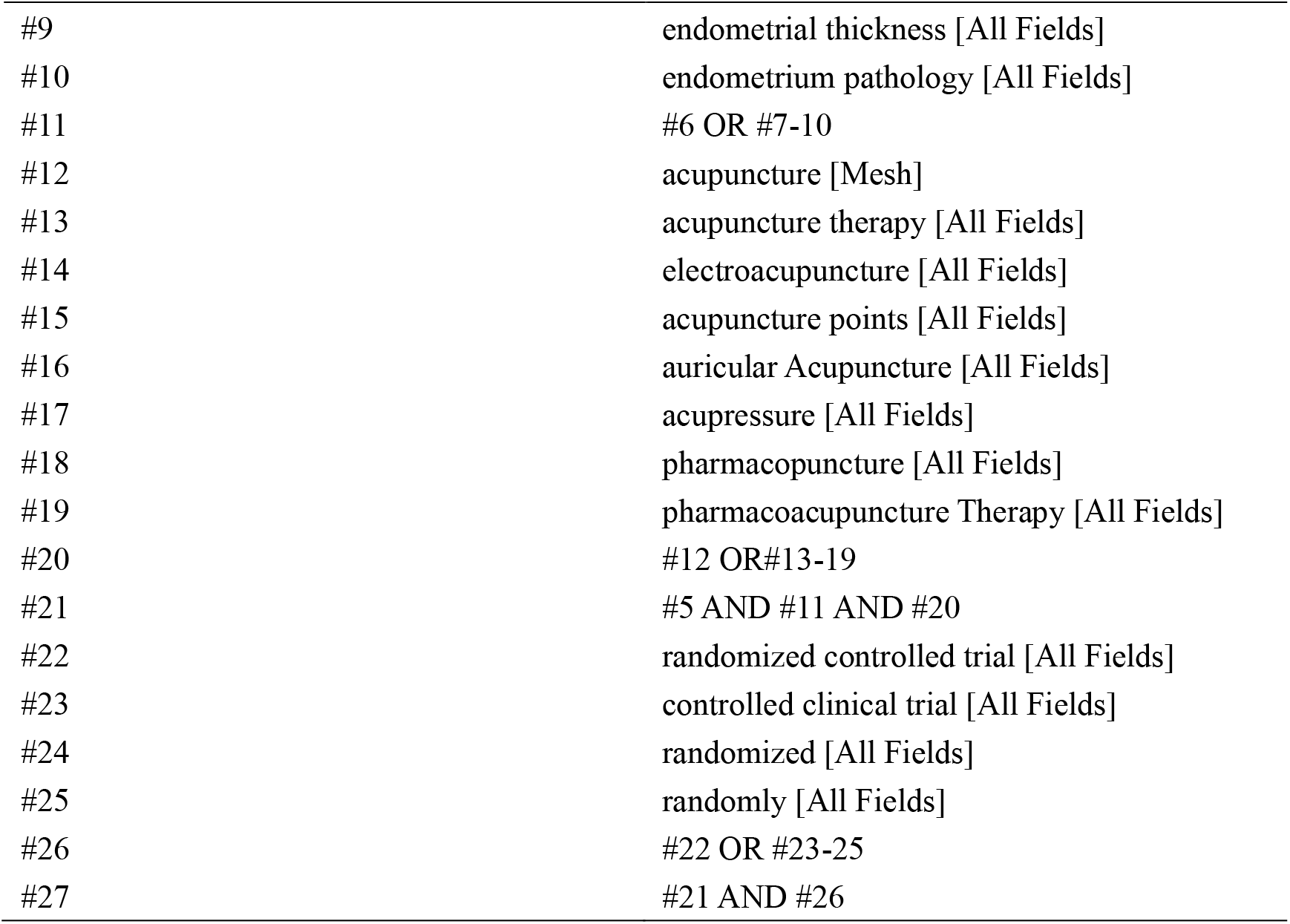
Search strategy used in PubMed database.

| Search Number | Search Items |
| --- | --- |
| #1 | infertility [All Fields] |
| #2 | infertile woman [All Fields] |
| #3 | Infertility epidemiology [All Fields] |
| #4 | Infertility therapy [All Fields] |
| #5 | #1 OR #2-4 |
| #6 | endometrial receptivity [Mesh] |
| #7 | uterine receptivity [All Fields] |
| #8 | thin endometrium [Mesh] |
| #9 | endometrial thickness [All Fields] |
| #10 | endometrium pathology [All Fields] |
| #11 | #6 OR #7-10 |
| #12 | acupuncture [Mesh] |
| #13 | acupuncture therapy [All Fields] |
| #14 | electroacupuncture [All Fields] |
| #15 | acupuncture points [All Fields] |
| #16 | auricular Acupuncture [All Fields] |
| #17 | acupressure [All Fields] |
| #18 | pharmacopuncture [All Fields] |
| #19 | pharmacoacupuncture Therapy [All Fields] |
| #20 | #12 OR #13-19 |
| #21 | #5 AND #11 AND #20 |
| #22 | randomized controlled trial [All Fields] |
| #23 | controlled clinical trial [All Fields] |
| #24 | randomized [All Fields] |
| #25 | randomly [All Fields] |
| #26 | #22 OR #23-25 |
| #27 | #21 AND #26 |

#### 2.3.2. Searching for other resources

For more experiments, we will search for lists of relevant references. Clinical trial registries, like Menstrual Disorders and Subfertility Group (MDSG) Specialized Register, Cochrane Central Register of Controlled Trials(CENTRAL), World Health Organization International Clinical Trials Registry Platform, Chinese clinical registry, and Clinical Trials. Gov. Manual searches of key journals and meetings such as European Society for Human Reproduction and Embryology (ESHRE) and American Society of Reproduction Medicine(ARSM) for relevant articles, including relevant journals and conference abstracts, by connecting with the coordinator.

### 2.4. Study selection and data extraction

Two reviewers will download the citations into NoteExpress software Version 2.6.1 (Aegean Sea software company, Beijing, China), and repeated research will be eliminated by software. Any disagreement will be resolved through further discussion with a third reviewer (FG.Wang). The selection process will be documented and summarized in a PRISMA flow chart **(Fig. 1)**.

**Figure 1.**
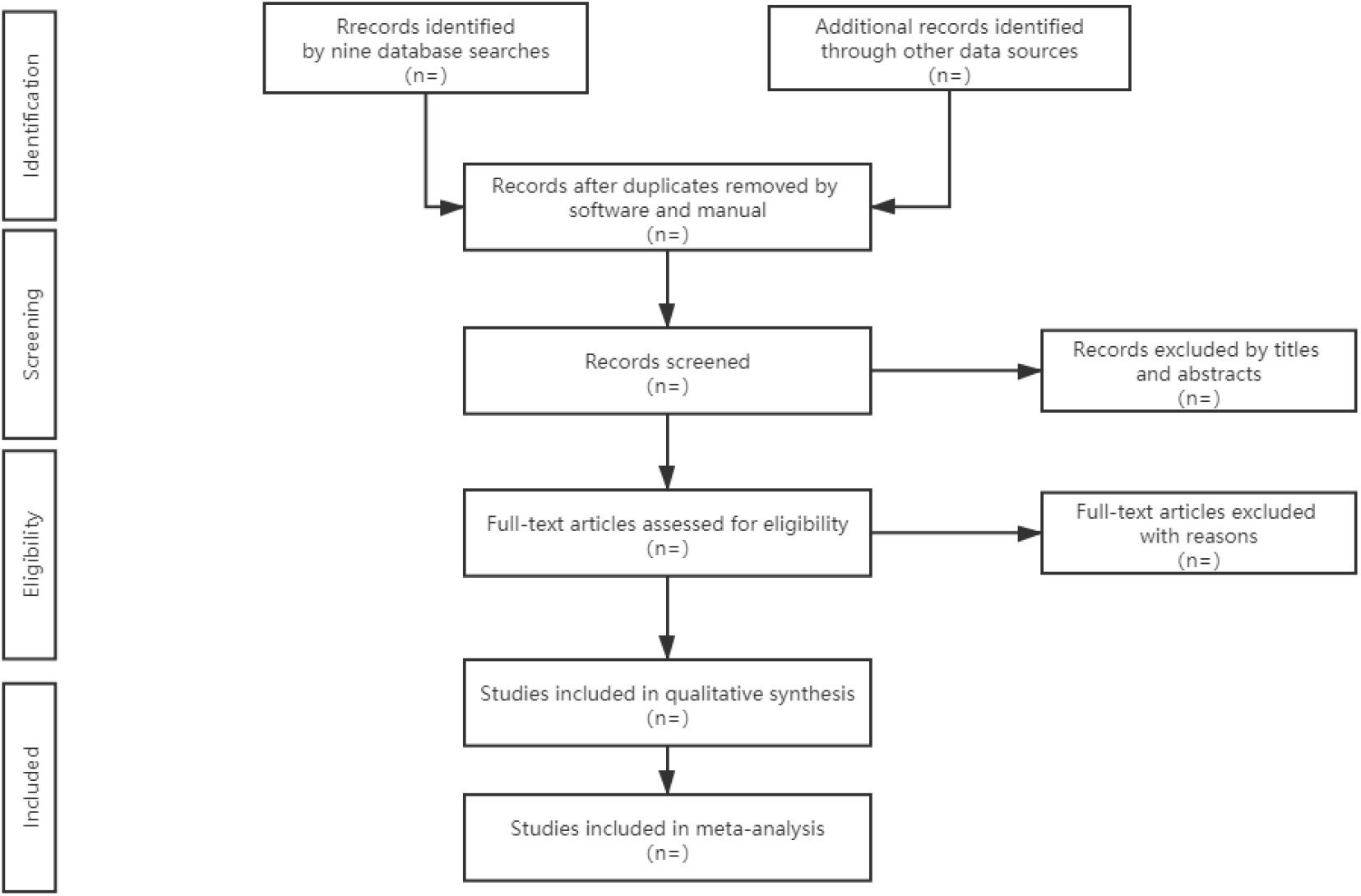
Flow diagram of study selection. This picture reflects the steps of research, selection, and explains the process of literature screening in detail.

All trial data extraction was performed independently by two review authors (XY.Zheng and HM. Yang). Data will be extracted onto a predeveloped sheet to assess the study characteristics (first author, publication year, sample size, duration of follow-up, number of participants, and finished numbers), the patient characteristics(mean age, body mass index(BMI), duration of infertility, the baseline of endometrial thickness), a dose of the interventions(the starting time of Acupuncture, the duration and frequency of Acupuncture sessions, and the time window of Acupuncture treatment), outcomes and reported adverse events. If the title or abstract appeared to meet the eligibility criteria, the full texts of the articles were obtained for further evaluation. If the data in a study were insufficient or ambiguous, one reviewer (XYZ) contacted the corresponding author by e-mail to get additional information.

### 2.5. Assessment of risk of bias in the included studies

Two review authors (XY.Zheng and HM.Yang) will independently assess the quality of the trails according to the PRISMA recommendations. The discrepancies were resolved by a third review author (FG, Wang) until consensus was reached. The risk of bias of RCTs will be assessed using the Cochrane Collaboration’s tool. The criteria consist of seven items: ① selection bias (random sequence generation and allocation concealment);② performance bias (blinding of participants and personnel); ③ detection bias (blinding of outcome assessment); ④ attrition bias (incomplete outcome data);⑤reporting bias (selective reporting); and ⑥other bias. Each study was evaluated as High, Low, or Unclear risk of bias for each item, and the assessment criteria were based on the Cochrane handbook^[38]^

### 2.6. Assessment of heterogeneity

The heterogeneity will be evaluated using both the I^2^ test and the P-value of the χ ^2^ test of heterogeneity. According to the Cochrane Handbook suggestions, we will use a fixed-effects model for data synthesis if the I^2^ value is <50%; otherwise, we will use a random-effects model for data synthesis.

### 2.7. Assessment of reporting biases

Publication bias will be tested by the visual inspection of the funnel plots. When a few studies are included in the analysis, the tests’ power is too low; therefore, publication bias was only examined if more than 10 study comparisons were included in the calculation.

### 2.8 Data synthesis

We will use the statistical package (RevMan5.4.0) provided by The Cochrane Collaboration to analyze data. For dichotomous data, such as CPR, endometrial pattern, and the rate of AEs, we expressed the results for each study as the risk ratios (RRs) with 95% confidence interval (CIs). And for the continuous data, such as endometrial thickness, clinical pregnancy rate, helical arterial blood flow index including resistive index (RI), pulse index (PI) peak systolic velocity/end-diastolic blood velocity (S/D), we expressed the results as the difference or standardized mean difference (SWD) with 95% CI. If data cannot be synthesized, we will use descriptive analysis to solve this problem.

### 2.9 Subgroup analysis

It is known that age is the most important indicator in infertility,which is highly relevant to reproductive ability.And also,different application time-point will lead to different effect in acupuncture treatment^[39]^,which is related to the sessions of acupuncture treatment.Therefore,We will conduct subgroups analysis as the following aspects:

1. clinical characteristics: mean age(≥ 35 or <35 years); duration of infertility (≥ 5 or <5 years)
2. different starting time (the different stage of the menstrual cycle or IVF) and the frequency and duration of Acupuncture treatment;
3. different types of Acupuncture treatment: manual Acupuncture, electro-acupuncture, TEAS, et al.;
4. different types of controls.

### 2.10. Sensitivity analysis

If there is pronounced heterogeneity between a group of studies, the reasons for the heterogeneity should be explored from multiple aspects, such as the characteristics of the research object and the degree of variation in intervention measures. We will assess the Effect of methodological quality, sample size, and missing data. If necessary, we will change the inclusion criteria (especially the controversial studies), exclude low-quality studies, use different statistical methods/models to analyze the same data.

### 2.11. Grading the quality of evidence

The included trials will be graded as low quality, high quality, or moderate quality based on the following criteria: (1) tests are considered low quality if either randomization or allocation concealment is assessed as a high risk of bias, regardless of the risk of other items; (2) trials are considered high quality when both randomization and allocation concealment will be assessed as a low risk of bias, and all other things are set the as low or unclear risk of bias in a trial; (3) tests were considered moderate quality if they did not meet criteria for high or low risk. Additionally, Assess the quality of evidence using the Grading of Recommendations Assessment, Development, and Evaluation (GRADE) framework; all judgments were fully described and presented in the risk of bias tables by the standards as below.

## 3. Discussion

The “China Infertility Survey Report” released in 2018 concluded that one-eighth of couples of childbearing age in the country face fertility problems, and the incidence becomes gradually younger. Female infertility is a complex disease with different pathogenic factors. Though the wide-spread use of the assisted reproductive technique (ART) has helped many women solve infertility problems, there is still lots of infertile patients suffering pregnancy outcome of low embryo implantation rate and high embryo loss rate due to low endometrium receptivity. Therefore, improving ER is the key to infertility and increase the IVF-ET pregnancy rate. Acupuncture has already become an excellent complementary therapy as an ancient therapy of traditional Chinese medicine, which improves outcomes of ART of women who were suffered from low endometrium receptivity. However, the results from randomized controlled trials were contradictory. Previous studies indicated that the effect of Acupuncture was dose-dependent. But a confirmative conclusion on Acupuncture for endometrium receptivity is rare, and the best dose of Acupuncture is not exact. Therefore, we will explore the relationship between the component of Acupuncture dose and endometrium receptivity. This study will collect evidence comprehensively, hoping to provide an evidence-based basis for Acupuncture treatment of female infertility and provide more useful information for medical staff and better advice for patients.

## 4. Patient and public involvement

No patients or the public participated in the preparation of the study protocol.

## 5. Ethics and dissemination

No ethical approval is required because the study is based on published data. The results of this systematic review will be disseminated through peer-reviewed publications.

## Data Availability

The datasets used or analysed during the current study are available from the corresponding author on reasonable request.

## Acknowledgements

We acknowledge professor Jie Yang, an outstanding Chengdu University of Traditional Chinese Medicine, Acupuncture and Tuina School, who helped us revise the manuscript.

## Contributors

Conceptualization: Hongmei YANG.

Data curation:Xianyan ZHENG, Hongmei YANG.

Formal analysis: Xianyan ZHENG, Hongmei YANG.

Investigation: Liying LIU, Hongmei YANG.

Methodology: Xianyan ZHENG, Liying LIU.

Project administration: Fangge WANG.

Resources: Liying LIU.

Software: Liying LIU, Hongmei YANG.

Supervision: Fangge WANG, Jinzhu HUANG.

Validation: Fangge WANG.

Visualization: Hang ZHOU.

Writing – original draft: Xianyan ZHENG, Hongmei YANG..

Writing – review & editing: Xianyan ZHENG.

## Patient and public involvement

Patients and/or the public were not involved in the design, or conduct, or reporting, or dissemination plans of this research.

## Patient consent for publication

Not required.

## Provenance and peer review

Not commissioned; externally peer reviewed

## Open access

This is an open access article distributed in accordance with the Creative Commons Attribution Non Commercial (CC BY-NC 4.0) license, which permits others to distribute, remix, adapt, build upon this work non-commercially,and license their derivative works on different terms, provided the original work is properly cited, appropriate credit is given, any changes made indicated, and the use is non-commercial. See: http://creativecommons.org/licenses/by-nc/4.0/.

## References

[1] Harper JC, Aittomaki K, Borry P, Cornel MC. Recent developments in genetics and medically assisted reproduction: from research to clinical applications 2018; 26(1):12–33.

[2] Roussos-Ross D, Rhoton-Vlasak AS, Baker KM, Arkerson BJ, Graham G. Casebased care for pre-existing or new-onset mood disorders in patients undergoing infertility therapy. J Assist Reprod Genet. 2018;35(8):1371–6

[3] Rostami Dovom M, Ramezani Tehrani F, Abedini M, et al. A population-based study on infertility and its influencing factors in four selected provinces in Iran (2008–2010). Iran J Reprod Med 2014;12:561–6.

[4] Ishihara O, Adamson D, Dyer S, Jd M, Nygren KG, Sullivan EA, ZegersHochschild F, Mansour R. International Committee for Monitoring Assisted Reproductive Technologies: world report on assisted reproductive technologies, Fertil Steril. 2015;103(2):402–13

[5] Opoien HK,Frdorcsak P,Omland AK, et al. In vitro fertilization is a successful treatment in rndometriosis- associated infertility. Frrtil Steril, 2012; 97 (4): 912–918.

[6] Pagidas K,Ying Y,Keefe D. Predictive value of pre-implantation genetic diagnosis for aneuploidy screening in repeated IVF-ET cycles among women with recurrent implantation failure. J Assist Reprod Genet, 2008;25(2-3):103–106.

[7] Simon A, Laufer N. Repeated implantation failure:clinical approach, Fertil Steril 2012;97(5):1039–43.

[8] Tan BK, Vandekerkhove P, Kennedy P, et al. Investigation and current management of recurrent IVF treatment failure in the UK. BLOG, 2005;112(6):773–780.

[9] Margalioth EJ, Ben-Chetrit A, Gal M. Investigation and treatment of repeated implantation failure following IVF-ET. Hum Reprod. 2006;21(12):3036–43.

[10] Diedrich K, Fauser B, Devroey P, et al. The role of endometrium and embryo in human implantation. HumReprod. 2007;13:365–377.

[11] Gao M, Sun Z, Zhao X, et al. Effect of different ovarian response after controlled ovarian hyperstimulation on pregnancy outcome of women undergoing in vitro fertilization. Journal of Shanghai Jiaotong University(Medical Science). 2013;1(33):33–45.

[12] Forman R, Fries N, Testart J, et al. Evidence for an adverse effect of elevated serum estradiol concentrations on embryo implantation. FertilSteril. 1988;49(1):118 –122.

[13] Pope CS, Cook EKD, Arny M, et al. Influence of embryo transfer depth on invitro fertilization and embryo transfer outcomes. FertilSteril. 2004;8(1):51–58.

[14] Lédée-Bataille N, Laprée-Delage G, Taupin JL, et al. Concent ration of leukemia inhibitory factor (LIF) in uterine flushing fluid is highly predictive of embryo implantation. Hum Reprod, 2002;17(1):213–218.

[15] Davis MC, Rozenwaks Z. Preparation of the Endometrium for Egg Donation. J Assist Reprod Genet. 1993;10:457–459.

[16] Gonen Y, Casper RF. Prediction of implantation by the sonographic appearance of the endometrium during controlled ovarian stimulation for in vitro fertilization(IVF). J A SSIT Reprod Genet. 2000;7(3):146–152.

[17] Yang JH, Wu MY, Chen CD, Jiang MC, Ho HN, Yang YS. Association of endometrial blood flow as determined by a modified color Doppler technique with subsequent outcome of in-vitro fertilization. Human reproduction (Oxford, England). 1999;14(6):1606

[18] Dentali F, Ageno W, Rezoagli E, et al. Low-dose aspirin for in vitro fertilization or intracytoplasmic sperm injection:a systematic review and a meta-analysis of the literature. J Thromb Haemost. 2012:10:2075–2085.

[19] Almog B, Shalom-Paz E, Dulort D, et al. Promoting implantation by local injury to the endometrium. Fertil Steri1. 2010;94:2026–2029.

[20] Lan VT, Khang VN, Nhu UH, et al. Atosiban improve implantation and pregnancy rates in patients with repeated implantation failure [J/OL]. Reprod Biomed Online. 2012;25:254–260.

[21] Potdar N, Uelbaya TA, Konje JC, et al. Adjunct low-molecular-weight heparin to improve live birth rate after recurrent implantation failure: a systematic review and meta-analysis. Hum Reprod Update. 2013;19:674–684.

[22] Uleicher N, Kim A, Michaeli T, et al. A pilot cohort study of granulocyte colony-stimulating factor in. 2013;28:172–177.

[23] Paulus WE, Zhang M, Strehler E, et al. Influence of Acupuncture on the pregnancy rate inpatients who undergo assisted reproduction therapy. Fertil Steril. 2002;77(4):721–724.

[24] Cai L, Li L, Zhao Y, Liu X, et al. Effects of Acupuncture method of strengthen spleen and invigorating kidney on ovulation and endometrial receptivity of infertile patients with PCOS in IVF-ET. SH. J. TCM, 2020;54(11):48–52

[25] Johnson D. Acupuncture prior to and at embryo transfer in an assisted conception unit-a case series. Acupuncture in medicine:journal of the British Medical Acupuncture Society. 2006;24(1):23–8.

[26] Zhang M, Huang G, Fe L. Effect of Acupuncture on pregnancy rate in embryo transfer. Zhongguo zhenjiu. 2002;22(8).

[27] Chen D, Chen S, Shi X. Clinical observation of Acupuncture for the treatment of multiple cystic ovary syndrome. Zhongguo zhenjiu. 2007;27(2):99–102.

[28] Wang X, Yuan H, Liu H, et al. Effect of warm needling on endometrial receptivity in kidney-yang deficiency patients with thin endometrium[J]. Shanehai J Acu-mox. 2019;38(7)

[29] Chen W, Liao J, Feng C, et al. Clinical Study on Endometrial Receptivity of Acupuncture and Medicine in Infertility[J]. Lishizhen medicine and meteria medica research. 2019;30(9):2180–2182

[30] Emily Wing Sze So, Ernest Hung Yu Ng, Yu Yeuk Wong, Estella Yee Lan Lau, William Shu Biu Yeung and Pak Chung Ho. A randomized double blind comparison of real and placebo acupuncture in IVF treatment[J]. Human Reproduction, 2009, 52(2):49–50.

[31] ZhongY, Zeng F, Liu W. et al.. Acupuncture in improving endometrial receptivity:a systematic review and meta-analysis. BMC Complement Altern Med, 2019;19(1):61

[32] Yang L, Lan Y, Yang L, et al. A meta-analysis of Acupuncture and moxibustion improving endometrial receptivity. Chinese archives of traditional chinese medicine, 2019;37(5):1102–1109

[33] Hu Q, He Y, Wang L, et al. Meta analysis on Acupuncture therapy for improving endometrial receptivity. Clinical journal of traditional Chinese medicine. 2017;29(1):61–67

[34] Juanjuan M A, Qinhua Z. Clinical study of acupuncture on improving endometrial receptivity, emotion of anxiety and depression, and pregnancy outcome in IVF-ET repeated implantation failure patients with syndrome of kidney deficiency and liver stagnation.

[35] Shamseer L, Moher D, Clarke M, et al. Preferred reporting items for systematic review and meta-analysis protocols(PRISMA-P) 2015:elaboration and explanation. BMJ (Clinical research ed) 2015;350: g7647.

[36] Kupesic S, Bekavac I, Bjelos D, Kurjak A. Assessment of endometrial receptivity by transvaginal color Doppler and three-dimensional power Doppler ultrasonography in patients undergoing in vitro fertilization procedures. J ultrasound in medicine : official journal of the American Institute of Ultrasound in Medicine. 2001;20(2):125–34.

[37] Yaman C, Mayer R. Three-dimensional ultrasound as a predictor of pregnancy in patients undergoing ART. J Turkish German Gynecological Assoc. 2012;13(2):128–34.

[38] Higgins JPT, Green S. Cochrane Handbook for Systematic Reviews of Interventions (version 5. 1. 0). Updated 2011. Available at: http://www.cochranehandbook.org. Accessed March, 2011.

[39] Wu R, Wang Y. Application time-point and effect observation of fire needling therapy in IVF-ET. Zhongguo Zhen Jiu. 2017 May 12;37(5):498–502

